# The overlap of genetic susceptibility to schizophrenia and cardiometabolic disease can be used to identify metabolically different groups of individuals

**DOI:** 10.1101/2020.06.23.20138271

**Authors:** Rona J. Strawbridge, Keira J. A. Johnston, Mark E. S. Bailey, Damiano Baldasarre, Breda Cullen, Per Eriksson, Ulf DeFaire, Amy Ferguson, Bruna Gigante, on behalf of the IMPROVE study, Philippe Giral, Nicholas Graham, Anders Hamsten, Steve E. Humphries, Sudhir Kurl, Donald M. Lyall, Laura M. Lyall, Matteo Pirro, Jill P. Pell, Kai Savonen, Bengt Sennblad, Andries J. Smit, Elena Tremoli, Tomi-Pekka Tomainen, Fabrizio Veglia, Joey Ward, Daniel J. Smith

**Affiliations:** Institute of Health and Wellbeing, University of Glasgow, Glasgow, UK; Health Data Research, UK; Department of Medicine Solna, Karolinska Institute, Stockholm, Sweden; Deanery of Molecular, Genetic and Population Health Sciences, College of Medicine and Veterinary Medicine, University of Edinburgh, Scotland, UK; School of Life Sciences, College of Medical, Veterinary & Life Sciences, University of Glasgow, Scotland, UK; Centro Cardiologico Monzino, IRCCS, Milan, Italy; Department of Medical Biotechnology and Translational Medicine, Università degli Studi di Milano, Milan, Italy; Cardiovascular Medicine Unit, Department of Medicine Solna, Karolinska Institutet, Stockholm, Sweden; Usher Institute, University of Edinburgh, UK; Assistance Publique - Hopitaux de Paris; Service Endocrinologie-Metabolisme, Groupe Hôpitalier Pitie-Salpetriere, Unités de Prévention Cardiovasculaire, Paris, France; Centre for Cardiovascular Genetics, Institute Cardiovascular Science, University College London, London, UK; Institute of Public Health and Clinical Nutrition, University of Eastern Finland, Kuopio, Finland; Internal Medicine, Angiology and Arteriosclerosis Diseases, Department of Clinical and Experimental Medicine, University of Perugia, Perugia, Italy; Foundation for Research in Health Exercise and Nutrition, Kuopio Research Institute of Exercise Medicine, Kuopio, Finland; Department of Clinical Physiology and Nuclear Medicine, Kuopio University Hospital, Kuopio, Finland; Department of Cell and Molecular Biology, National Bioinformatics Infrastructure Sweden, Science for Life Laboratory, Uppsala University, Uppsala, Sweden; Department of Medicine, University Medical Center Groningen and University of Groningen, Groningen, The Netherlands; Public Health and Clinical Nutrition, Department of Medicine, University of Eastern Finland, Kupiou, Fnland

**Keywords:** Genetics, Schizophrenia, type 2 diabetes, blood pressure, hypertension, cardiovascular disease, obesity, clustering

## Abstract

Understanding why individuals with severe mental illness (Schizophrenia, Bipolar Disorder and Major Depressive Disorder) have increased risk of cardiometabolic disease (including obesity, type 2 diabetes and cardiovascular disease), and identifying those at highest risk of cardiometabolic disease are important priority areas for researcher. We explored whether genetic variation could identify individuals with different metabolic profiles. Loci previously associated with schizophrenia, bipolar disorder and major depressive disorder were identified from literature and those overlapping loci genotyped on the Illumina CardioMetabo and Immuno chips (representing cardiometabolic processes and diseases) were selected. In the IMPROVE study (high cardiovascular risk) and UK Biobank (general population) multidimensional scaling was applied to genetic variants implicated in both mental and cardiometabolic illness. Visual inspection of the resulting plots used to identify distinct clusters. Differences between clusters were assessed using chi-squared and Kruskall-Wallis tests. In IMPROVE, genetic loci associated with both cardiometabolic disease and schizophrenia (but not bipolar or major depressive disorders) identified three groups of individuals with distinct metabolic profiles. The grouping was replicated in UK Biobank, albeit with less distinction between metabolic profiles. This study provides proof of concept that common biology underlying mental and physical illness can identify subsets of individuals with different cardiometabolic profiles.

## Introduction

Individuals with serious mental illness (such as schizophrenia (SCZ), major depressive disorder (MDD) and bipolar disorder (BD)) have a reduced life expectancy (10-15years for BD, 15-20years for SCZ [1]). This is likely due to the well-established increased prevalence of cardiovascular and metabolic disorders compared to the general population. For example, obesity is up to 3.5-fold higher in those with SCZ [2], type 2 diabetes is ∼2-fold higher in those with MDD, BD or SCZ [2], and cerebrovascular disease is increased by up to 3.3-fold in those with BD [2]. Understanding this increased risk and identifying individuals at highest risk of metabolic and cardiovascular disease are important priority areas for researchers and healthcare providers.

Historically, the increased risk and prevalence of cardiometabolic disease (CMD) has been attributed to social determinants and lifestyle factors (including poor diet, sedentary behaviour, alcohol and substance use) that co-exist with serious mental illness and effects of psychotropic medication [2], however there is growing evidence that there might be common biological mechanisms underlying both mental and psychiatric illness. As genetic data is stable over an individual’s lifetime, and not influenced by disease course, genetic approaches are ideal for investigation of common biology in comorbid conditions. The identification of genetic variants robustly associated with a wide range of psychiatric and cardiometabolic phenotypes by international genetics consortia has enabled the exploration of relationships between psychiatric and cardiometabolic conditions.

Genome-wide genetic correlations between psychiatric and cardiometabolic traits provide evidence for underlying common biology. Correlations have been described between depression and obesity (rg=0.12) or cardiovascular disease (rg=0.42) [3]. Evidence of causal relationships between psychiatric and cardiometabolic traits have also been described [1, 4, 5]. However, the mechanisms involved have yet to be uncovered and therefore this knowledge has had no clinical impact.

Here we tested whether a novel approach using multi-dimensional scaling (MDS) of genetic variation associated with psychiatric and cardiometabolic disorders could aid stratification of individuals into groups with differing cardiometabolic risk profiles.

## Results

### The IMPROVE and UK Biobank studies

The demographic characteristics of the IMPROVE, UK Biobank subsets 1 (UKB1) and 2 (UKB2) are provided in Table 1. At baseline, individuals in IMPROVE (a European high cardiovascular-risk cohort) were older, more overweight and more likely to have T2D, hypertension or medication for hypertension or lipid-lowering medication than the UKB subsets (white British general population cohort). UKB1 and UKB2 were very similar, with lower frequency of hypertension at follow-up in UKB1 (51.5%) compared to UKB2 (62.0%) but slightly larger carotid Intima-media thickness (cIMT, indicative of vessel wall remodelling) measures in UKB2 to UKB1. Despite different proportions of UKB1 and UKB2 completing the mental health questionnaire, the frequencies of BD, MDD and GAD were similar.

**Table 1:**
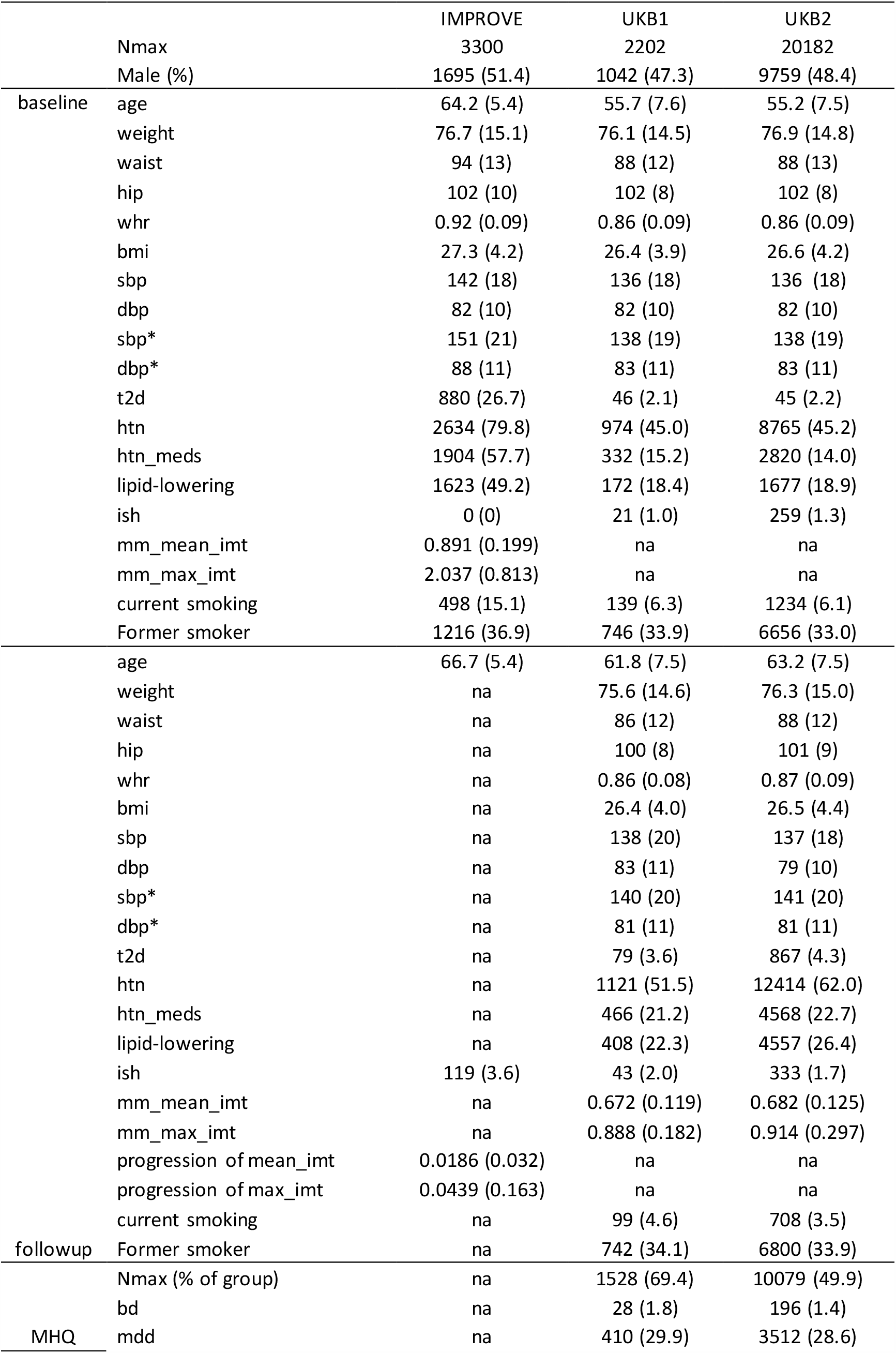
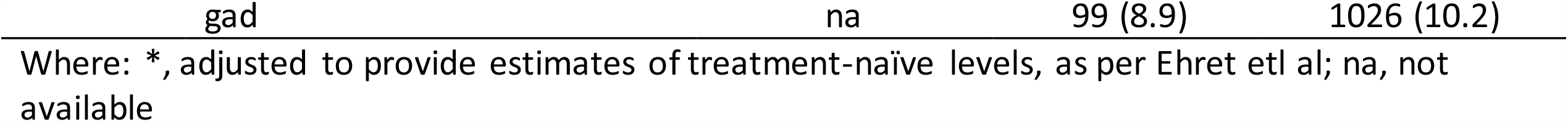
Demographic characteristics of IMPROVE and UKB participants

Figure 1 provides a schematic overview of the analysis procedure.

**Figure 1:**
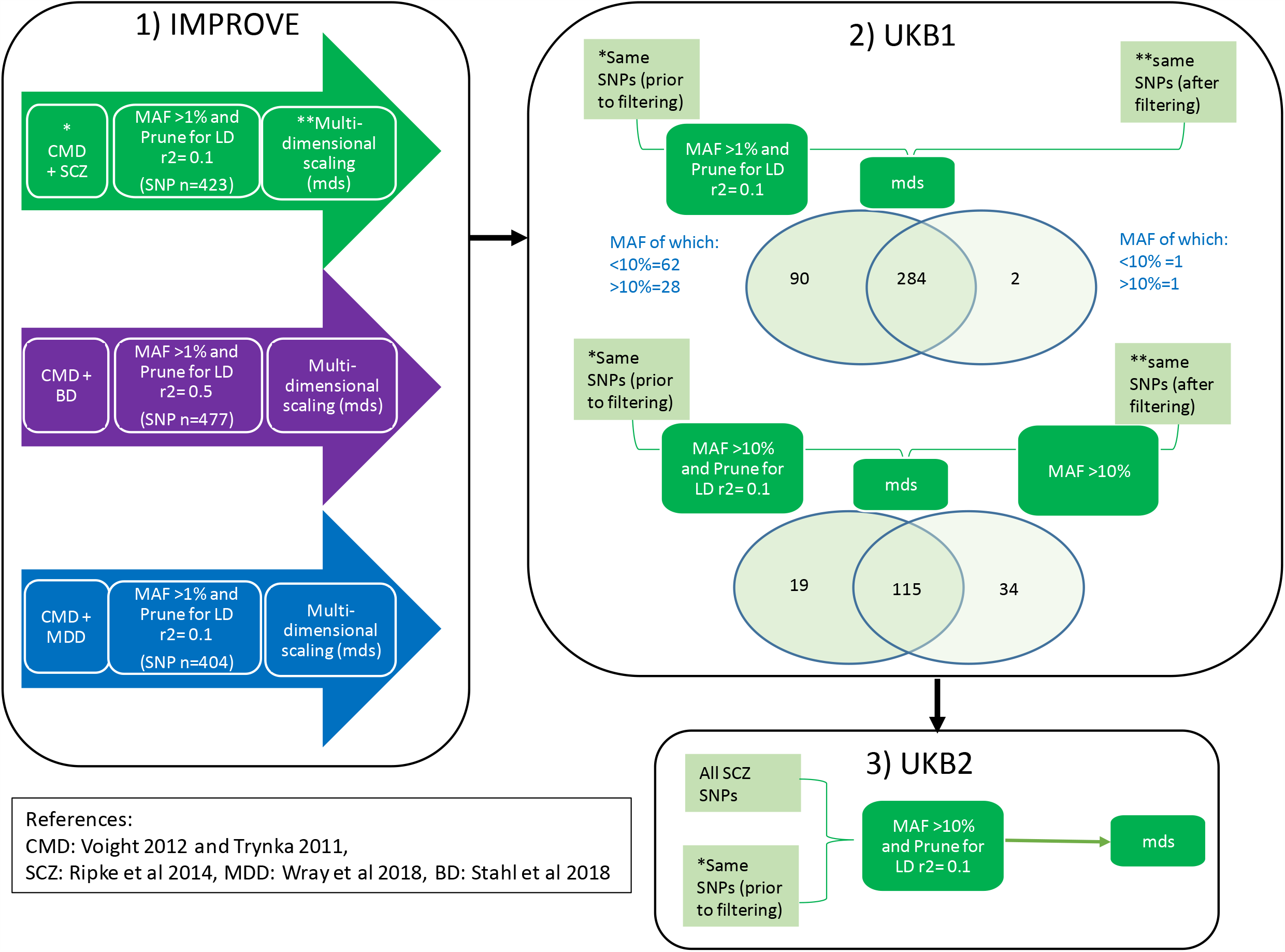
Schematic of the analysis procedure used to identify clusters.

### SCZ-CM loci can identify metabolically distinct groups of individuals in IMPROVE

When using IMPROVE and single nucleotide polymorphisms (SNPs) with a minor allele frequency (MAF) >1%, implicated in both SCZ and CMD (SCZ-CMD), plotting the first two multi-dimensional scaling components (C1 and C2) demonstrated 3 groups of individuals (by visual inspection) (Figure 2a). Separation was predominantly due to C1, and whilst C1 is nominally significantly correlated with latitude (rho=-0.036, p=0.0339), the clustering is not being driven by latitude (SFigure 1). SNPs with MAF as low as 1% might differ across populations (even within the same ancestry grouping), therefore robustness to MAF threshold also assessed. When using MAF >5% showed additional groups (Figure 2b), whereas MAF >10% showed similar groups to MAF >1. Assignment to groups was consistent using MAF >1% and MAF >10% (STable 1). The three groups appear to have modest differences in cardiometabolic profiles (Table 2): Group 3 had a significantly lower frequency of hypertension (P=0.004) and lower fastest progression of cIMT (P=0.002). This is surprising given the (non-significant) higher rates of smoking in this group. Group 2 had (non-significantly) lower rates of T2D than the other groups.

**Table 2:**
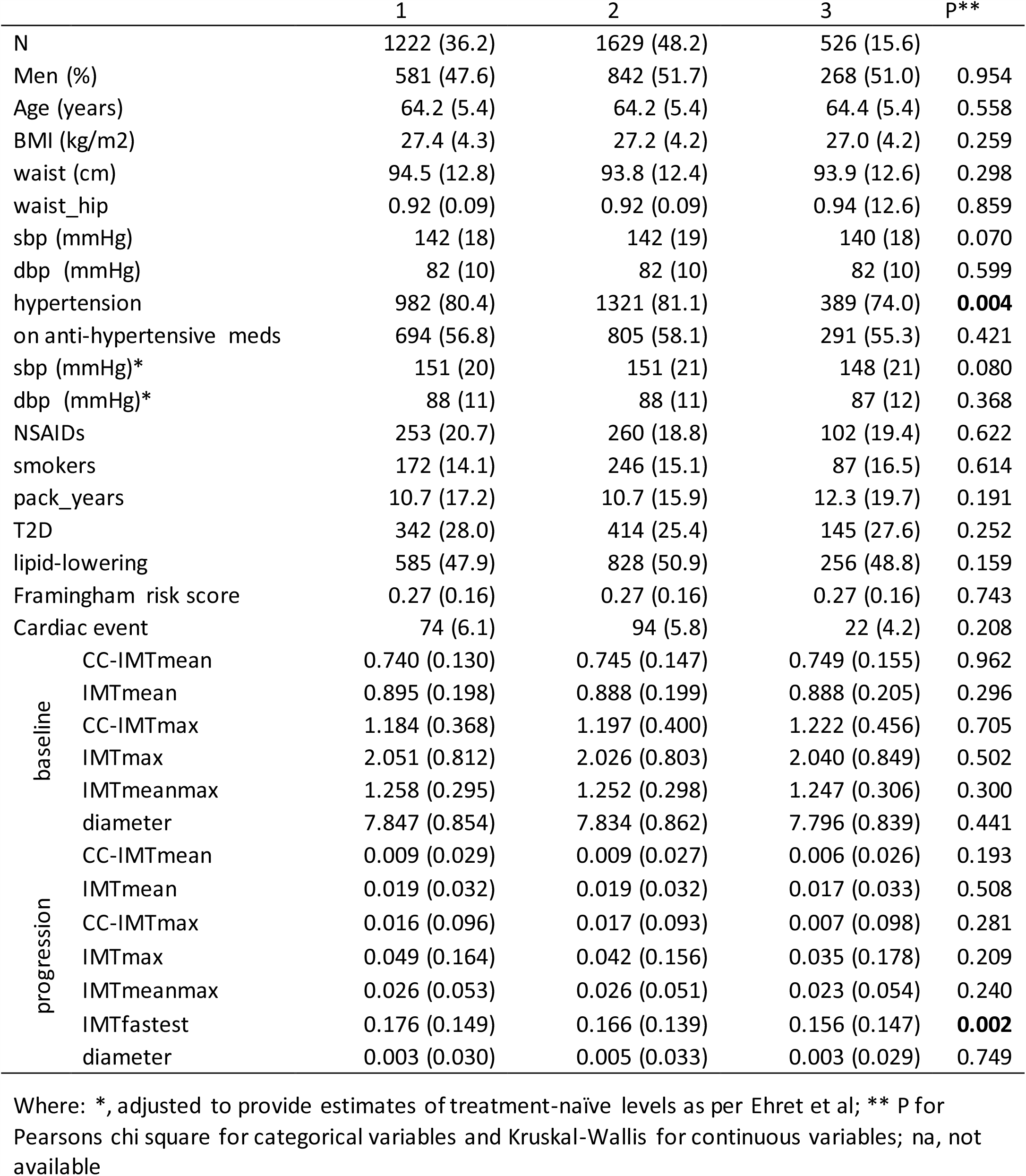
Demographic characteristics of the IMPROVE participants, by cluster (MAF>10%)

**Figure 2:**
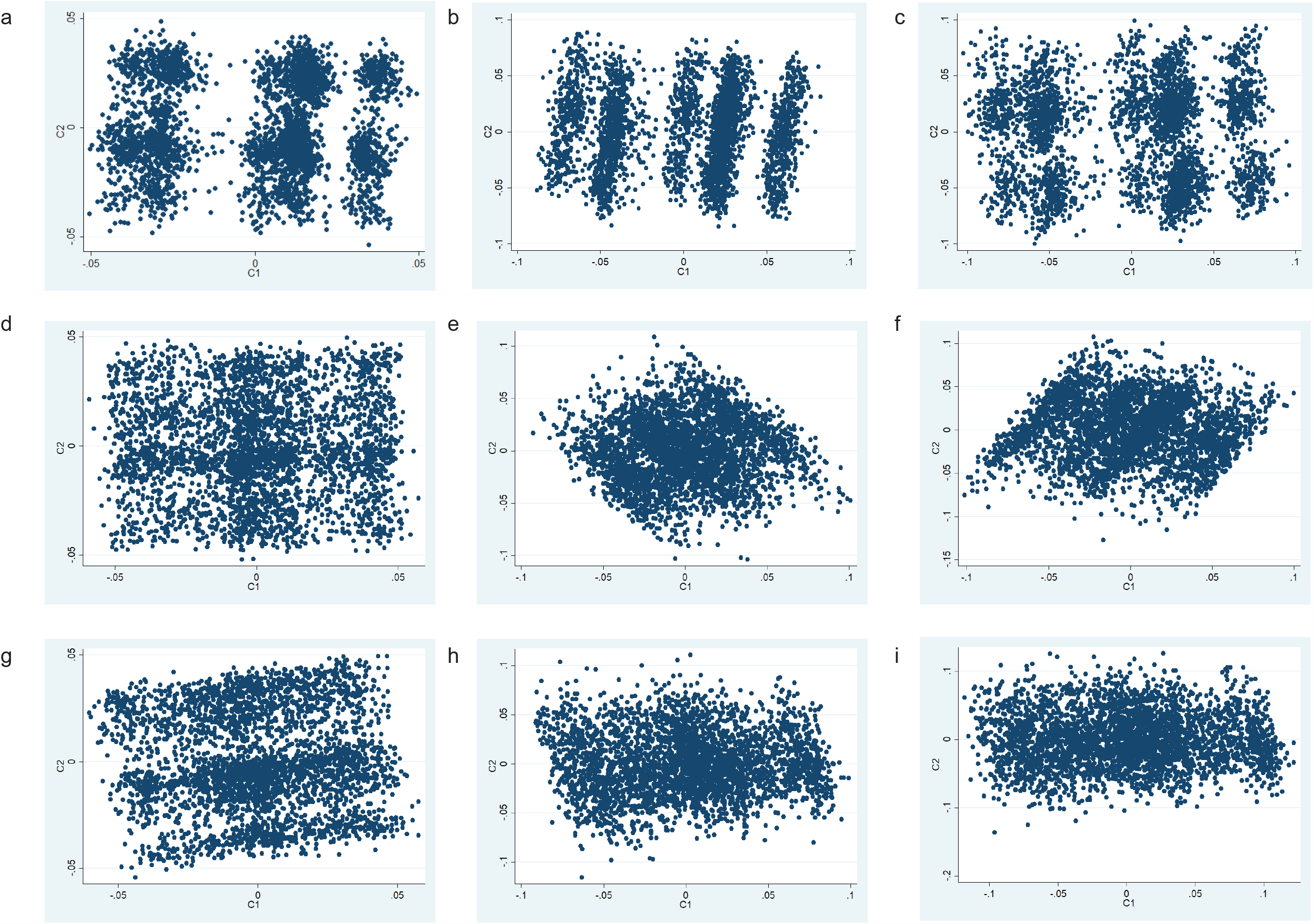
Results of MDS analysis in IMPROVE, using the loci in common between CMD and SCZ with MAF >1%, b) MAF >5% or c) MAF >10%; CMD and MDD with d) MAF >1%, e) MAF >5% or f) MAF >10%; CMD and BD with g) MAF >1%, h) MAF >5% or i) MAF >10%. Each data point is an individual therefore the individuals who are closer together are more genetically similar.

No separation into groups was observed when using MDD-CM SNPs, irrespective of the MAF filter used (Figure 2d-f). For BD-CM SNPs (Figure 2g-i), grouping is apparent at MAF >1%, but not when MAF >5% or 10% were considered.

### Validation of method and sensitivity testing of clustering in UKB1

In order to assess whether MDS analysis of SCZ-CM SNPs could reproducibly identify three groups of individuals, validation of the method was attempted in UKB1. Firstly, to directly replicate the analysis conducted in IMPROVE (Figure 3a and b), the post-filtering SNPs from IMPROVE were used (Figure 3c), however the grouping is not convincing as there is little separation between the groups. Secondly, to assess robustness of the method to differences in MAF and LD structure between populations, the SCZ-CM SNPs were filtered for MAF and LD in UKB1. As noted in Figure 1, the majority of SNPs included in the two approaches were the same. Unsurprisingly, the SNPs that differed were mainly those with MAF<10%. Using SCZ-CM and conducting MAF and LD filtering in UKB1, nine groups are evident when using SNPs with MAF >1% (Figure 3d), whereas three groups are observed when using SNPs with MAF >10% (Figure 3e). When comparing the metabolic profiles of the 3 groups, no significant differences were seen (Figure 4a and Table 3). This is unsurprising, given that it is a smaller cohort with a lower cardiovascular burden.

**Table 3:**
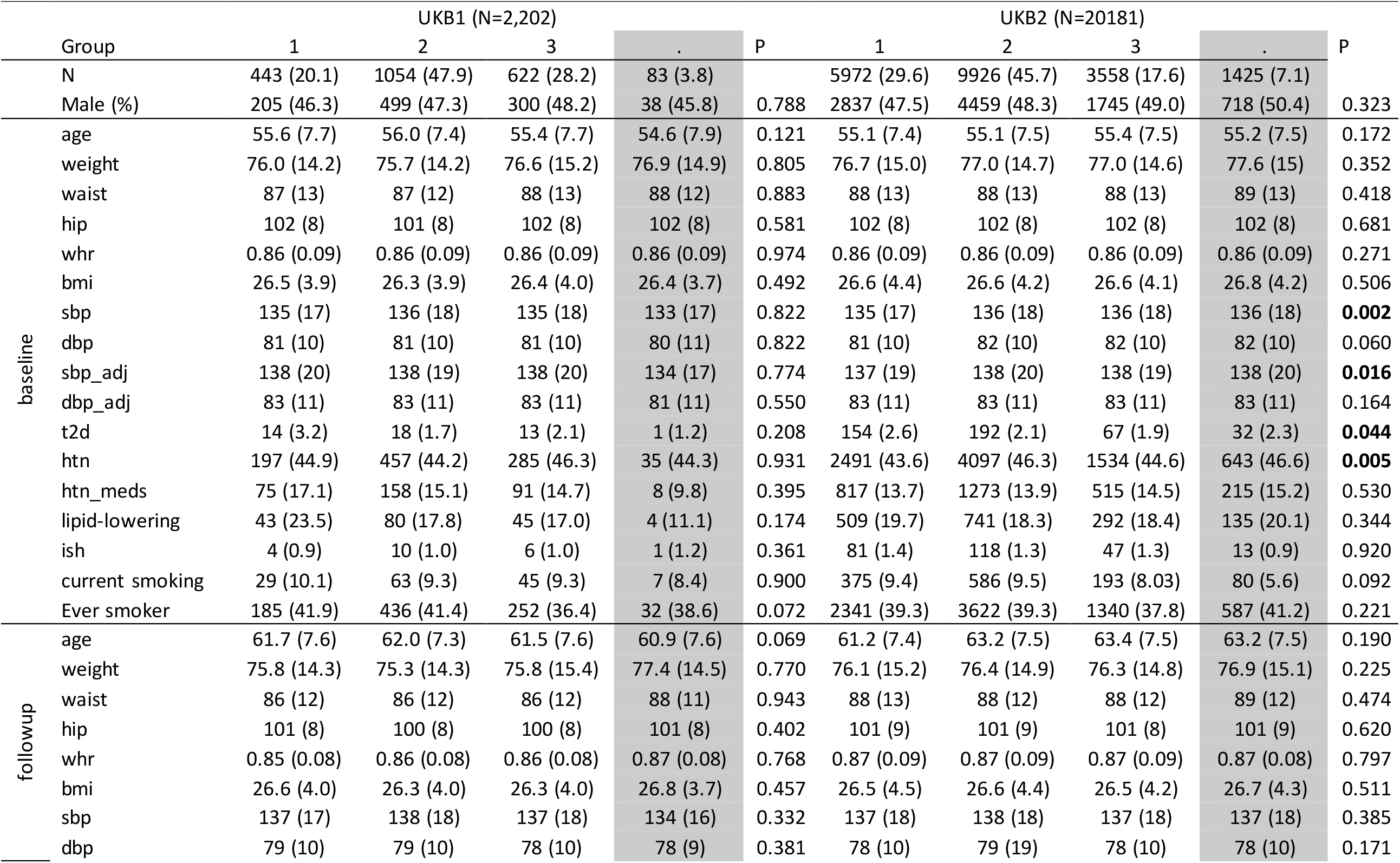
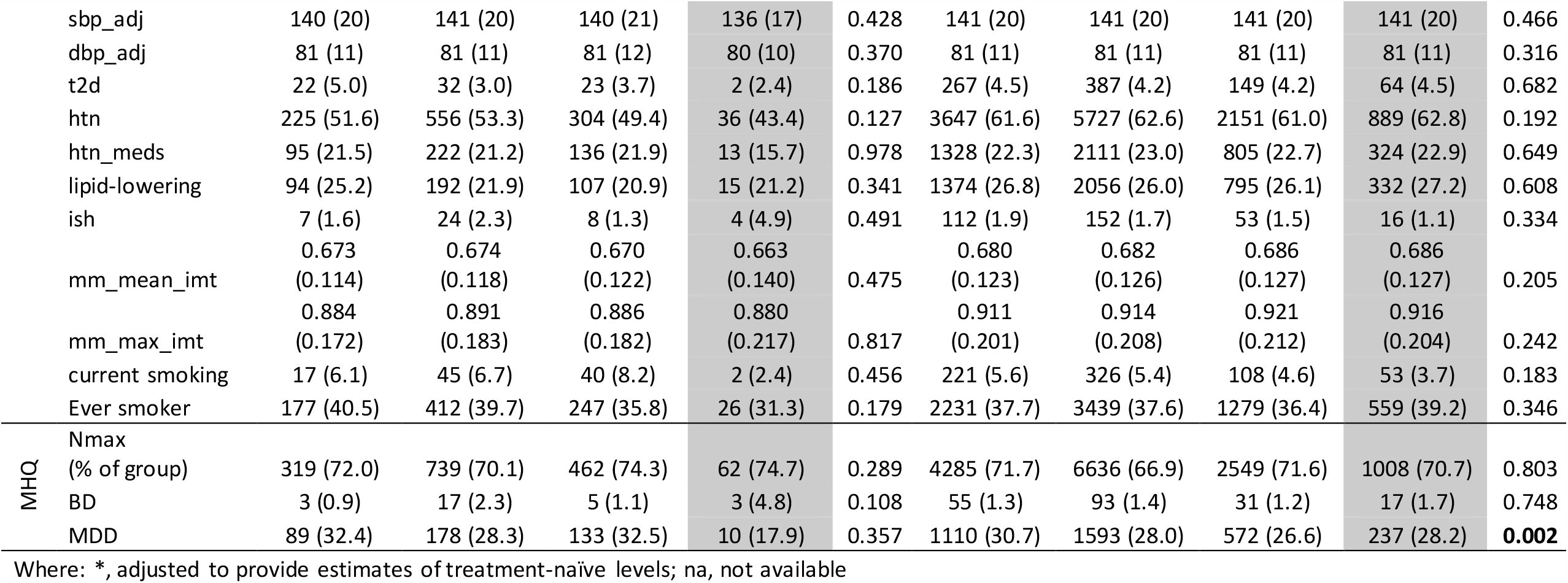
Demographic characteristics of the UKB1 and UKB2 participants, by cluster

**Figure 3:**
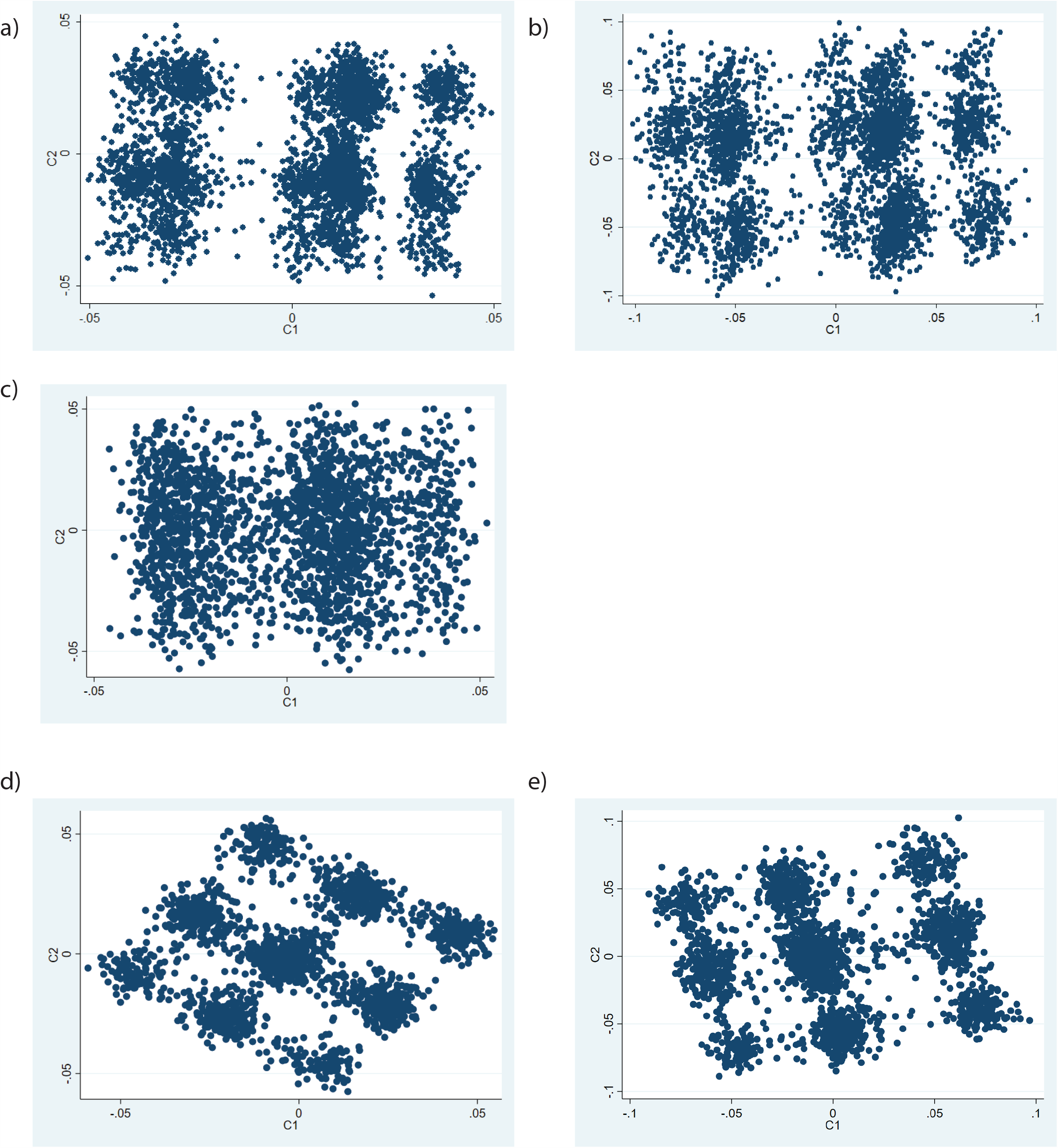
Sensitivity testing in UKB1. For comparison, IMPROVE MDS analysis using a) MAF >1% and MAF >10%. MDS analysis in UKB1 using c) the same post-filtering SNPs as for IMPROVE, d) the same pre-filtering SNPs with MAF >1% in UKB1 and e) the same pre-filtering SNPs with MAF >10% in UKB1. Each data point is an individual therefore the individuals who are closer together are more genetically similar.

**Figure 4:**
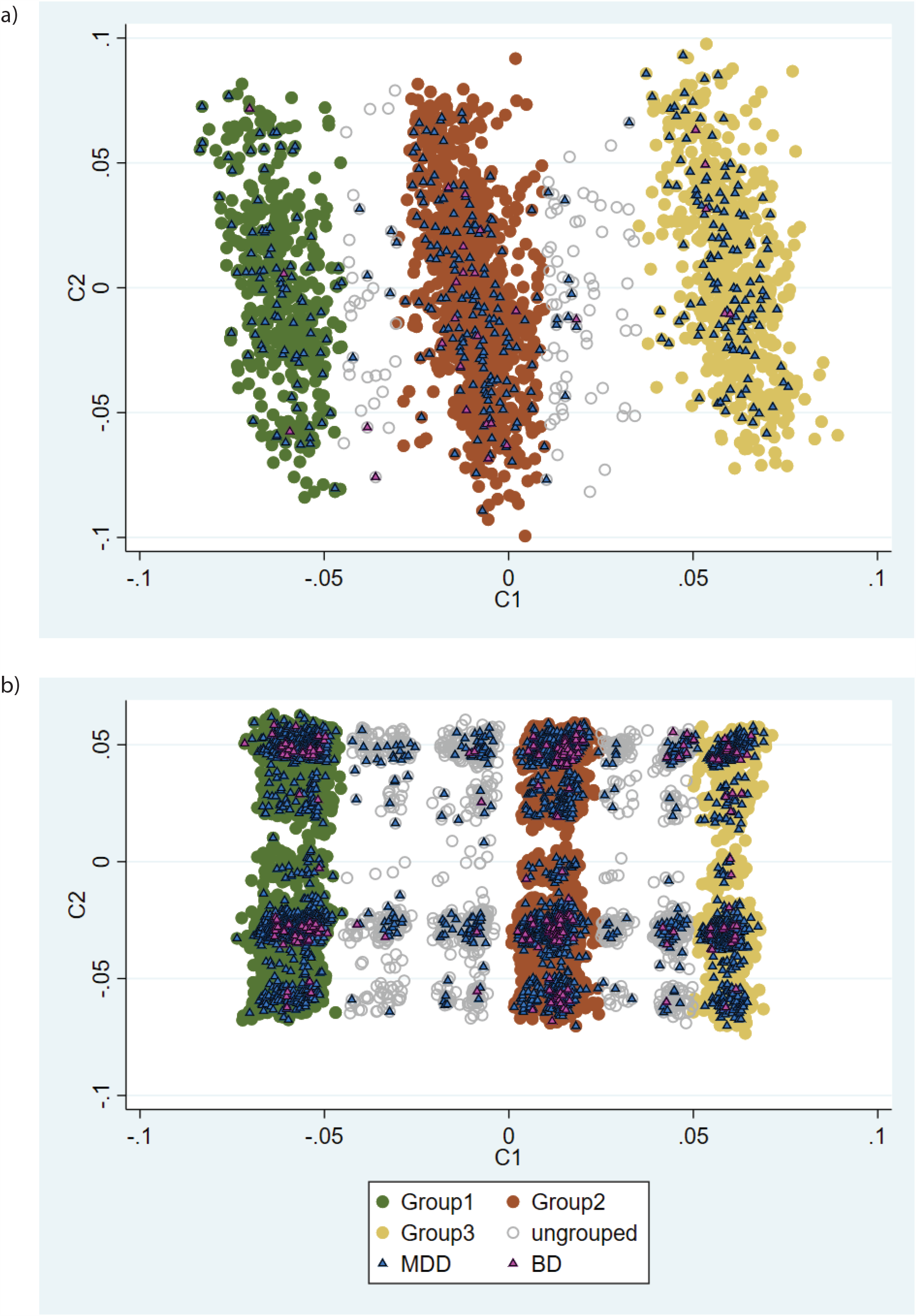
Comparison of the three clusters identified in a) UKB1 and b) UKB2 (lower panel). Each data point is an individual therefore the individuals who are closer together are more genetically similar.

### Validation of metabolic differences between clusters in UKB2

In an attempt to replicate the clustering and validate the metabolic differences between groups, the larger UKB2 subset was analysed. As filtering with MAF >10% and 1% gave similar clusters, filtering with MAF >10% was applied as it is more likely to generalise to other populations. Again, three major groups were identified (Figure 4b), similar to those identified in IMPROVE and UKB1. Additional clusters between the major three groups were apparent, but they account for ∼7% of the studied population, and were omitted from the groups.

Significant (but clinically modest) differences were observed in baseline SBP, SDP adjusted for blood-pressure medication, and frequency of hypertension and T2D (Table3). These effects were not observed at follow-up, potentially due to lifestyle or medications changes in response to baseline observations. It was also noted that the frequency of MDD but not BD differed between the groups. The number of SCZ in UKB2 is too low to provide meaningful statistics.

### Impact of MDD/BD on clusters

As phenotypes and genetic loci for SCZ overlap with those for MDD and BD, it is perhaps unsurprising to see that the clusters include different proportions of individuals with MDD. To investigate whether these individuals were driving the clustering, the process was repeated in those without BD/MDD separately from those with these diagnoses (using SNPs with MAF >10%). In those without mental illness, similar to the overall UKB2, there were there main groups, intermediate clusters accounting for 7.4% of the sample (Figure 5a). In those with mental illness the three clusters were observed, with better between-group separation and only 1.3% of the sample being ungrouped (Figure 5b). Small but significant differences between groups were observed for blood pressure measures and rates of hypertension, in both those with and without mental illness (STable 2). These results suggest that this method is applicable to the general population, as well as those with increased genetic burden for mental illness,

**Figure 5:**
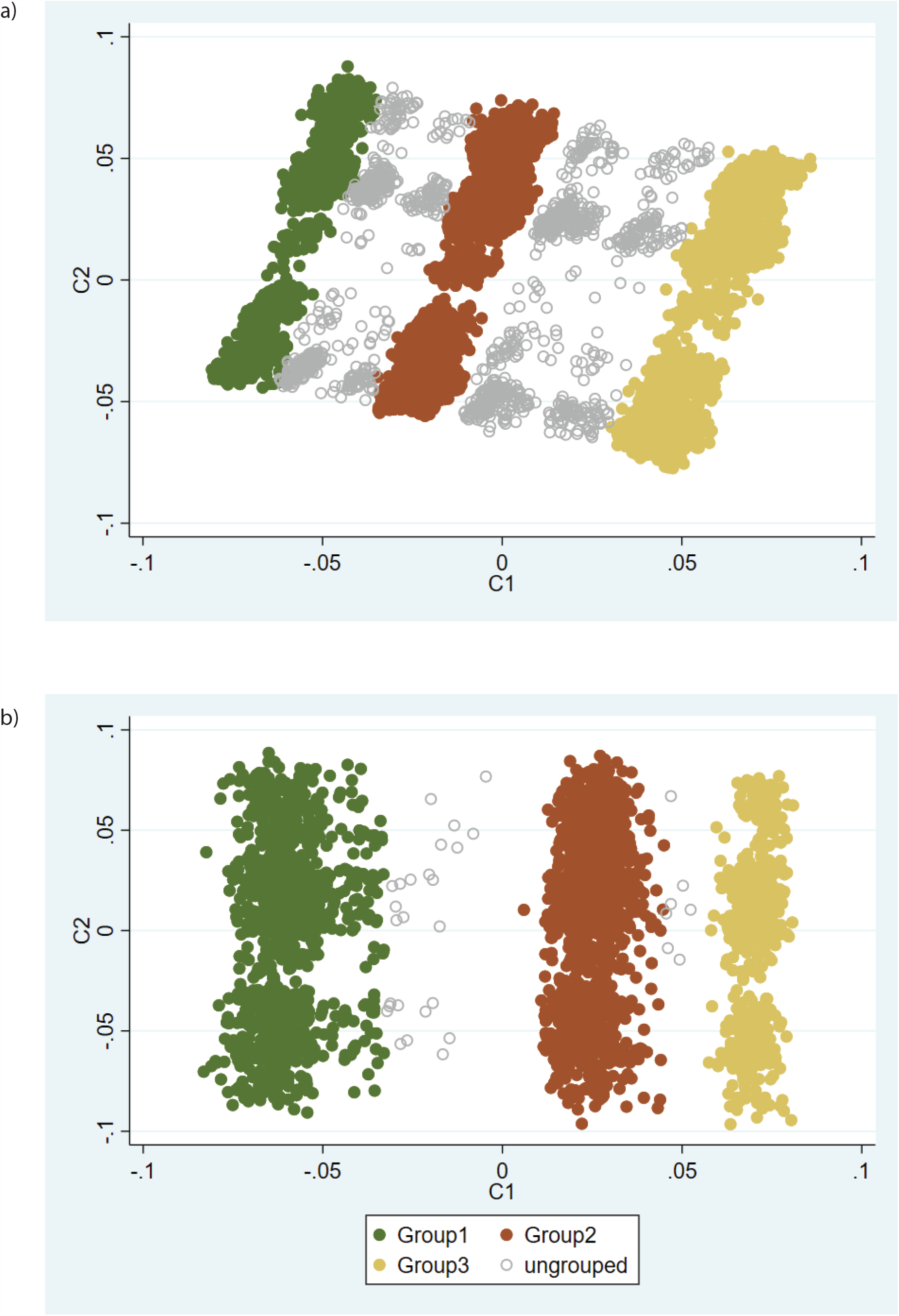
Comparison of the three clusters identified in UKB2 in individuals a) without and b) with mental illness. Each data point is an individual therefore the individuals who are closer together are more genetically similar.

### All genetic loci associated with SCZ do not identify clusters in UKB

To determine whether it is common biology (ie. Overlap in loci for SCZ and CMD) *per se*, rather than SCZ in general that drives the clustering, the same procedure was followed using all SNPs in loci associated with SCZ in UKB2, with the same MAF and LD filtering being applied prior to MDS analysis. As shown in SFigure 2, SNPs in loci associated with SCZ do not separate individuals into groups, although nine closely grouped clusters are visible when plotting MDS components 3 and 4. This confirms that it is the overlap of SCZ and CMD loci, and therefore probably common biological mechanisms, which are driving the clustering.

## Discussion

This study provides proof of principle that, using the genetic overlap between SCZ and cardiometabolic disorders, subsets of individuals with different metabolic profiles can be identified. These findings support the existence of mechanisms common to SCZ and blood pressure regulation.

The discovery cohort IMPROVE deliberately recruited to identify genes and biomarkers associated with the risk of cardiovascular diseases, at a time when psychiatric disorders were typically excluded from non-psychiatric studies, therefore only a portion of the spectrum of psychiatric genetic burden is represented. In contrast, UKB1 and UKB2 are general population cohorts and therefore have a wider spectrum of both psychiatric and cardiometabolic disorder genetic burden, although it is recognised that the recruitment skews this distribution towards to the healthier segment of the population [6]. It is therefore both striking that the grouping was present in IMPROVE, and unsurprising that the blood pressure and hypertension differences between groups were more modest in UKB2 than those in IMPROVE.

That the metabolic profiles of the groups did not completely agree between the 3 cohorts was disappointing, however the repeated observation of between-group differences in T2D and blood pressure/hypertension deserves further attention. If the method can be refined to better identify whether an individual is at increased risk of either hypertension or T2D would be of immense value. Even if the method is only robust in high CMD-risk populations (such as those with family history, multiple risk factors or psychiatric diagnoses), it would be of clinical importance.

It is interesting that the analyses using BD and MDD genetic loci did not enable clustering of individuals in the same way as was observed for SCZ, particularly given that BD and SCZ demonstrate an overlap in genetic loci. There are several possible explanations for this, most notably the ability to identify genetic loci for each mental illness: SCZ is clinically a more severe phenotype with diagnostic criteria that are relatively specific (for example psychotic episodes). In comparison, MDD spans a wide spectrum severity, with phenotypic heterogeneity potentially diluting or obscuring some true genetic effects. Whilst BD can be considered an intermediate (some symptoms more severe than MDD, most are less severe than for SCZ) diagnostic criteria for MDD and BD overlap to a large degree as both involve episodes of depression, meaning that there is potential for misdiagnosis and therefore dilution of genetic effects for either trait. Another explanation is that the mechanisms leading to CMD in SCZ differ from those in MDD or BD, with processes that are represented on the CardioMetabo and Immuno chips failing to capture some pathological mechanisms. With this in mind, the finding of different frequencies of MDD in the groups was not anticipated, as the MDD genetics did not achieve any form of grouping, and the overlap of MDD and SCZ genetics is modest. However, MDD is highly heterogeneous, therefore it would be of interest to further explore whether there are any differences between the MDD cases in each group, specifically whether any of the groups corresponds to the recently proposed atypical depression subtype [7, 8].

Genetic correlation analyses have begun to explore the common biology and causal relationships between psychiatric and cardiometabolic diseases [1, 3, 8], however these methods assume that the entire genome influences both sets of traits. The small to moderate correlations could suggest that it is only a portion of the genome that has common effects. In contrast, the current study focuses on only the parts of the genome that have been implicated in both psychiatric and CMD. Whilst this study does not bring us any closer to understanding the mechanisms underlying the common pathological mechanisms, it does suggest that exploration of the SCZ-CMD loci could have clinical utility, irrespective of mechanistic understanding.

One limitation is that these analyses were conducted in individuals of European ancestry and as SNPs were filtered by MAF and linkage disequilibrium, it is not possible to generalise them to other populations. Another limitation is that the CardioMetabo and Immuno chips do not include all loci implicated in cardiometabolic disorders. Since these chips were described (2012 and 2011 respectively), many more loci involved in many more processes have been identified. However, as more and more samples are available for GWAS analyses, loci are being identified with smaller and smaller effect sizes. Therefore whilst not all possible information is captured by using the CardioMetabo and Immuno chips, the loci with the largest effects are represented.

In conclusion, this study provides proof of concept that common biology underlying mental and physical illness is probable and can distinguish subsets of individuals with differing metabolic profiles, even if full understanding of mechanisms is lacking. Given that large-scale genotyping is not available to healthcare providers, there is currently limited potential for translation of this into clinical practice. Further investigation with longitudinal datasets, particularly in high CVD risk populations, would define whether or not there is potential for clinical value in this method.

## Methods

### Cohorts: phenotyping and genotyping

<u>The IMPROVE study</u> has been described previously [9, 10]. In short, 3700 individuals aged between 54-79 years with high CVD risk profiles (the presence of at least 3 classical CVD risk factors, including family history of CVD, type 2 diabetes, hypertension, hyperlipidaemia and smoking) were recruited from seven centres in Finland, Sweden, the Netherlands, France and Italy. At baseline, individuals completed lifestyle and medical questionnaires and anthropometric measures taken. Blood was sampled for DNA extraction and clinical biochemistry and stored for further biochemical analyses. Detailed ultra-sound examination of the carotid intima-media thickness (cIMT) was conducted at baseline, 15 months and 30 months. Linear regression using all data points was used to calculate progression of cIMT. Mental illness was not assessed, however it is believed that if there is mental illness in this cohort it is likely to be subclinical. All participants provided written informed consent and the study was conducted in accordance with the Helsinki Declaration. Ethical approval was granted by the regional ethics borad for each recruitment centre.

The IMPROVE study was genotyped on the Illumina Cardio-Metabo [11] and Immuno chips [12], therefore cardiometabolic disorders (including immune and inflammatory components) were well represented. Standard quality control procedures were conducted, namely exclusion of SNPs for low call rate (<95%) and deviation from Hardy-Weinberg Equilibrium (p< 1×10^−6^) and exclusion of samples for low call rate (<95%), sex-mismatch, cryptic relatedness. Quality control was conducted on each chip separately, followed by a further round of quality control on the combined chip.

<u>The UK Biobank</u> (UKB) has been described previously [13, 14]. Approximately 500,000 volunteers aged 39-73 years were recruited from 22 centres across the UK. At baseline, detailed questionnaires on sociodemographic factors, lifestyle factors and medical history were completed by all individuals. Measurements of anthropometric variables were recorded and blood samples were taken for DNA extraction. Subsequently (4-8 years after baseline), subsets of participants were invited for follow-up measurements and extensive imaging. All participants provided written informed consent and ethical approval was granted by the NHS national Research Ethics Service. This work was conducted under projects #6533 (Smith) and #1755 (Pell).

Ultrasound measurement of cIMT was conducted in a pilot phase of ∼2500 individuals (henceforth denoted as UKB1) followed by a subsequent phase including ∼22,000 individuals (denoted UKB2) using the same recruitment and measurement protocol. cIMT measurements were generally consistent with the measurements available in IMPROVE. A mental health/thoughts and feelings questionnaire was also completed by a subset of participants, which enabled estimation of life history of MDD and BD. For both UKB1 and UKB2, 73% of participants completed the mental health questionnaire.

Genome-wide genotyping was conducted and standard quality control procedures were applied by the UK Biobank team [15]. Imputation was conducted using the Haplotype reference consortium and 1000 Genomes with standard pre- and post-imputation quality controls being applied by the UK Biobank team (further information is provided in [15-17]).

### Multi-dimensional scaling (MDS) to identify clusters

Genome-wide genetic loci reported to be associated with SCZ [18], MDD [19] and BD [20] were identified. SNPs within these (SCZ, MDD or BD) loci which were present on the CardioMetabo and Immuno chips were selected [11, 12] (denoted SCZ-CM SNPs, MDD-CM SNPs or BD-CM SNPs, respectively). SNPs with MAF >1% were included (Stable 3). A schematic diagram of the analyses steps is provided in Figure 1.

In IMPROVE, each set of SNPs (SCZ-CM SNPs, MDD-CM SNPs or BD-CM SNPs) were pruned by pairwise LD (parameters 50, 5, 0.1) using PLINK [21]. Individuals with >1% missing genetic data were excluded prior to clustering. Clustering was performed using multi-dimensional scaling, implemented in PLINK, using default settings. Multidimensional scaling essentially measures similarity between individuals, in this case using the patterns of genetic variation as the assessment criteria [22, 23]. Individuals with similar genetic sequences are deemed more similar to each other than those with less similar genetic sequences.

Subsequently in UKB1, SCZ-cardiometabolic SNPs only were used and individuals with >1% missing genetic data were excluded prior to clustering. MDS analyses was conducted using either exactly the same SNPs as were used in IMPROVE (ie SCZ-CM SNPs after filtering for MAF and LD in IMPROVE) or SCZ-CM SNPs with filtering for MAF and LD being done in UKB1.

Finally, in UKB2, Individuals with >1% missing genetic data were excluded prior to clustering. MDS analysis was conducted on SCZ-CM SNPs with filtering for MAF and LD in UKB2, or on all SCZ SNPs after MAF filtering and pruning in UKB2.

The first two MDS components (C1 and C2) were plotted for visual assessment. Ideally a negative control experiment would be included, however as current evidence suggests that most genetic variants are highly pleiotropic and that complex traits overlap with each other to a large degree. Therefore choosing a “negative control” trait is not straightforward.

### Statistical analyses

In IMPROVE, Spearmans rank correlation coefficients were used to assess the relationship between the MDS components and latitude. For IMPROVE, UKB1 and UKB2, Differences between groups were assessed by Pearsons chi squared test for categorical values and Kruskal-Wallis test for continuous variables. All statistical analyses were conducted in Stata (version 11.0). The threshold for significance was set at p<0.05. No adjustment for multiple testing was applied, because these analyses are exploratory rather than definitive and secondly because most of the cardiometabolic phenotypes tested are interrelated and thus are not independent tests.

## Data Availability

The datasets generated during and/or analysed during the current study are available from the corresponding author on reasonable request.

## Acknowledgements

The authors thank all participants and staff of the IMPROVE and UK Biobank studies. This work uses data provided by patients and collected by the NHS as part of their care and support. <u>IMPROVE</u> was supported by the European Commission (Contract number: QLG1-CT-2002-00896), the Swedish Heart-Lung Foundation, the Swedish Research Council (projects 8691 and 09533), the Knut and Alice Wallenberg Foundation, the Foundation for Strategic Research, the Stockholm County Council (project 592229), the Strategic Cardiovascular and Diabetes Programmes of Karolinska Institutet and Stockholm County Council, the European Union Framework Programme 7 (FP7/2007-2013) for the Innovative Medicine Initiative under grant agreement n° IMI/115006 (the SUMMIT consortium), the Academy of Finland (Grant #110413), the British Heart Foundation (RG2008/08, RG2008/014) and the Italian Ministry of Health (Ricerca Corrente). The <u>UK Biobank</u> was established by the Wellcome Trust, Medical Research Council, Department of Health, Scottish Government and Northwest Regional Development Agency. UK Biobank has also had funding from the Welsh Assembly Government and the British Heart Foundation. Data collection was funded by UK Biobank.

RoJS is supported by a UKRI Innovation-HDR-UK Fellowship (MR/S003061/1). LML is supported by the JMAS Sim Fellowship for depression research from the Royal College of Physicians of Edinburgh. AF is supported by an MRC Doctoral Training Programme Studentship at the University of Glasgow (MR/K501335/1). KJAJ is supported by an MRC Doctoral Training Programme Studentship at the Universities of Glasgow and Edinburgh. DJS acknowledges the support of a Lister Prize Fellowship (173096) and MRC Mental Health Data Pathfinder Award (MC_PC_17217).

## Author Contributions

Study conception, design, data analysis (RJS) or acquisition (DB, UdF, AH, SHE, TPT), interpretation (RJS, KJAJ, PE, BG, SEH, DML, LML, BS, DJS), manuscript preparation or revision (RJS, KJAJ, MESB, DB, BC, PE, UdF, AF, BG, PG, NG, AH, SEH, SK, DML, LML, MP, JPP, KS, BS, AJS, TPT, FV, JW, DJS).

## Additional Information

The authors have no competing interests to declare.

